# FACTORS ASSOCIATED WITH ADHERENCE OF HEALTH WORKERS TO HAND HYGIENE PRACTICES AT LUBAGA HOSPITAL, KAMPALA CITY

**DOI:** 10.64898/2026.07.29.26359261

**Authors:** Nambi Flavia, Kizito Omona

**Affiliations:** Department of Health Services Management, Faculty of Health sciences, Uganda Martyrs University, Kampala-Uganda

**Keywords:** Adherence to Hand Hygiene Practice, Hand Hygiene Practices, Individual-level Factors, Institutional or Organizational Factors

## Abstract

**Background:** Hand hygiene is the most effective and cost-efficient measure for preventing healthcare-associated infections (HAIs). However, adherence among healthcare workers remains suboptimal, particularly in low-resource settings.

**Purpose/Aim:** This study assessed the level of adherence to hand hygiene practices and the factors associated with adherence among healthcare workers at Lubaga Hospital, Kampala City.

**Methods:** An analytical cross-sectional study was conducted among 216 healthcare workers selected using stratified random sampling. Data were collected through direct observation using the WHO “Five Moments for Hand Hygiene” checklist and a structured, self-administered questionnaire. Data were analyzed using SPSS version 20. Descriptive statistics were used to summarize the data, while inferential analysis included Pearson’s Chi-square tests and Modified Poisson regression to estimate crude and adjusted prevalence ratios (cPR and aPR) with 95% confidence intervals at a significance level of p ≤ 0.05.

**Results:** Overall adherence to hand hygiene practices was 69.9%, while 30.1% of participants were non-adherent. At the individual level, gender, cadre, and attitude toward hand hygiene were significantly associated with adherence. Male healthcare workers were less likely to adhere compared to females (aPR = 0.490, 95% CI: 0.226– 0.969), while nurses and midwives were more than twice as likely to adhere compared to other cadres (aPR = 2.213, 95% CI: 1.113–4.398). Participants with a positive attitude toward hand hygiene were also significantly more likely to adhere (aPR = 1.462, 95% CI: 1.227–3.226). Resource-related factors, including the availability of alcohol-based hand rub and timely replenishment of supplies, were not significantly associated with adherence. Organizational factors such as recent training in hand hygiene and infection prevention (aPR = 1.771, 95% CI: 1.673–2.989), presence of reminders (aPR = 1.747, 95% CI: 1.538–5.949), feedback on performance (aPR = 0.339, 95% CI: 0.156–0.738), and teamwork (aPR = 3.006, 95% CI: 1.424–6.348) were significantly associated with improved adherence.

**Conclusion:** Adherence to hand hygiene practices among healthcare workers was moderate but remains suboptimal. Behavioral and organizational factors, particularly training, feedback, teamwork, and attitude, play a mo**re** significant role in influencing and adherence than resource availability alone.

## Introduction

According to the United States Centers for Disease Control and Prevention (CDC), hand hygiene involves cleaning hands with soap and water, alcohol-based hand rub (ABHR), or surgical antisepsis to remove or kill microorganisms and prevent infection transmission in healthcare settings [1]. It is the most effective and cost-efficient measure for preventing healthcare-associated infections (HAIs), yet global adherence among health workers remains suboptimal, particularly in low- and middle-income countries [2]. In Uganda, persistent gaps in hand hygiene compliance have been linked to limited infrastructure, heavy workloads, inadequate supplies, and inconsistent availability of ABHR at points of care [3,4].

Although quality improvement initiatives have enhanced knowledge and compliance, significant barriers (including workflow constraints, resource limitations, and behavioral factors) remain in many facilities in Uganda [5]. Understanding these factors is critical for strengthening infection prevention and control efforts. Therefore, this study investigated the factors that are associated with the adherence of health workers to hand hygiene practices at Lubaga Hospital, Kampala City.

## Materials and methods

### Study design and study setting

This study employed a quantitative analytical cross-sectional research design to assess factors associated with health workers’ adherence to hand hygiene practices at Lubaga Hospital, Kampala District. A cross-sectional design is appropriate because it allows for the collection of data at a single point in time, enabling the measurement of the current level of hand hygiene adherence and examination of the relationship between adherence and the independent variables (individual-level factors, availability of hand hygiene resources, and institutional/organizational determinants). This design is justified for several reasons, namely, its ability to examine multiple factors simultaneously/concurrently [6], thereby providing an efficient and evidence-aligned method for examining the multifactorial determinants of hand hygiene adherence at Lubaga Hospital.

This study was conducted at Lubaga Hospital, also known as Uganda Martyrs’ Hospital Lubaga, a major private not-for-profit, community hospital located on Lubaga Hill in Lubaga Division, in the western part of Kampala District, Uganda’s capital city. The hospital is one of the oldest and most established health facilities in the country, having been founded in 1899 by the Missionary Sisters of Our Lady of Africa, and is recognized as the oldest Catholic hospital in Uganda. Lubaga Hospital is owned by the Roman Catholic Archdiocese of Kampala and is accredited by the Uganda Catholic Medical Bureau (UCMB), functioning as both a referral and teaching hospital for the region. Geographically, Lubaga Hospital is situated approximately 5.5 km southwest of Mulago National Referral Hospital and about 5 km west of Kampala’s central business district, making it easily accessible to a large urban population. The hospital sits adjacent to the historic Lubaga Cathedral, further anchoring it within an important religious and social hub on Lubaga Hill. The coordinates of the facility are 0°18′15.0″N, 32°33′10.0″E, at an elevation of approximately 1,249 meters above sea level

### Study population and selection criteria

The study population consisted of health workers employed at Lubaga Hospital, a major private, not-for-profit community hospital located on Lubaga Hill in Lubaga Division, Kampala City. The target population included all categories of health workers who are directly involved in patient care or who handle clinical materials. These included medical officers, clinical officers, nurses, midwives, laboratory personnel, pharmacists, and infection control staff. Health workers in such roles routinely interact with patients, handle equipment, and perform procedures that require strict adherence to the World Health Organization’s “Five Moments for Hand Hygiene.” Given the hospital’s sizeable workforce and its broad patient care activities, spanning outpatient care, inpatient care, emergency services, surgical theatres, maternity, pediatrics, renal services, and diagnostics, the study population provides a robust and contextually relevant sample for examining how individual, resource-related, and institutional factors associated with adherence to hand hygiene practices.

The study included Health workers who had worked at the hospital for at least one month, allowing adequate exposure to institutional IPC practices, workflows, and hand hygiene resources, and those who consented to participate in the study. These criteria ensure that participants have sufficient experience with the hospital’s hand hygiene systems, resource availability, and organizational environment.

### Sample size and sampling method

The sample size was determined using the Krejcie and Morgan formula for estimating a single population proportion [7]. In the absence of a facility-specific estimate of hand hygiene adherence at Lubaga Hospital, a conservative prevalence of 50% was adopted to maximize the sample size and enhance representativeness. Using a 95% confidence level and a 5% margin of error, an initial sample size of 384 participants was obtained. Given that the population of eligible health workers at Lubaga Hospital is finite, the sample size was adjusted using the finite population correction (FPC) formula. Based on the estimated number of eligible health workers within the hospital, the adjusted sample size was 216 participants. This sample was considered adequate to assess the factors associated with health workers’ adherence to hand hygiene practices. The study employed a stratified random sampling technique to select 216 health workers from Lubaga Hospital. Stratification was used to ensure proportional representation of the different professional cadres and clinical departments involved in patient care. This approach minimized sampling bias and enhanced the representativeness of the sample, thereby improving the validity of findings on factors associated with hand hygiene adherence among health workers.

#### Data Collection

Data were collected using two complementary tools: a WHO Hand Hygiene Direct Observation Checklist and a structured self-administered questionnaire. These tools were selected to assess health workers’ adherence to hand hygiene practices and the factors associated with adherence.

A standardized WHO Hand Hygiene Direct Observation Checklist was used to determine the level of adherence to hand hygiene practices among health workers. The tool is based on the WHO Five Moments for Hand Hygiene and was used to record hand hygiene opportunities and corresponding actions during routine patient care activities.

A structured self-administered questionnaire was used to collect data on individual and institutional factors associated with hand hygiene adherence. The questionnaire assessed knowledge, attitudes, training, workload, perceptions of hand hygiene, leadership support, supervision, and the availability of hand hygiene resources. Data collection was conducted in four phases: training of research assistants, participant recruitment, direct observation, and questionnaire administration.

Prior to data collection, research assistants received training on the study objectives, data collection procedures, research ethics, and the use of study tools to ensure consistency and reliability. Eligible health workers were then approached, informed about the study, and invited to participate after providing written informed consent. Following consent, participants were observed during routine clinical practice using the WHO Hand Hygiene Direct Observation Checklist. Observations were conducted discreetly across different departments and work shifts to minimize observation bias. Immediately after the observation period, participants completed a structured self-administered questionnaire in a private setting to obtain information on factors associated with hand hygiene adherence.

#### Data Analysis

Data were entered, cleaned, and analyzed using Statistical Package for Social Sciences (SPSS) version 20. Descriptive statistics, including frequencies, percentages, means, and standard deviations, were used to summarize participants’ characteristics and study variables. Findings were presented using tables, figures, and narrative summaries.

Inferential analysis was conducted to examine factors associated with adherence to hand hygiene practices. Pearson’s Chi-square test was initially used to assess associations between the dependent and independent variables. Given that hand hygiene adherence was a common outcome, Modified Poisson regression with robust error variance was employed to estimate prevalence ratios, which provide a more appropriate measure of association than odds ratios for cross-sectional studies with common outcomes [8,9]. Variables that were statistically significant at the bivariate level (p ≤ 0.05) were included in the multivariable model. Crude prevalence ratios (cPRs) and adjusted prevalence ratios (aPRs) with corresponding 95% confidence intervals were calculated to identify factors independently associated with hand hygiene adherence. Statistical significance was considered at a p-value of ≤ 0.05.

## Results

### Background Characteristics of Respondents

A total of 216 respondents participated in the study. The majority of participants were aged below 40 years (184, 85.2%), while only 32 (14.8%) were aged 40 years and above. Most respondents were female (182, 84.3%), with males representing a much smaller proportion (34, 15.7%). Regarding professional cadre, over half of the participants were in nursing (115, 53.2%), followed by others (69, 31.9%) and midwifery (32, 14.8%). Participants were drawn from various units of work. The largest proportion worked in the theatre (56, 25.9%), followed by the maternity ward (42, 19.4%), the outpatient department (OPD) (36, 16.7%), the nursing department (26, 12.0%), and the mother-to-child transmission (MTCT) area (25, 11.6%). Smaller proportions were from the emergency ward (12, 5.6%), postnatal ward (11, 5.1%), and children’s ward (8, 3.7%) (Table 1).

**Table 1:** Background Characteristics of Respondents.

| Characteristic | Frequency (N = 216) | Percent % |
| --- | --- | --- |
| **Age** |  |  |
| <40 Years | 184 | 85.2 |
| 40 Years and above | 32 | 14.8 |
| **Gender** |  |  |
| Male | 34 | 15.7 |
| Female | 182 | 84.3 |
| **Cadre** |  |  |
| Nursing | 115 | 53.2 |
| Midwifery | 32 | 14.8 |
| Others | 69 | 31.9 |
| **Unit of work** |  |  |
| Nursing Department | 26 | 12 |
| Theatre | 56 | 25.9 |
| Maternity Ward | 42 | 19.4 |
| MTCT Area | 25 | 11.6 |
| OPD | 36 | 16.7 |
| Postnatal Ward | 11 | 5.1 |
| Children's Ward | 8 | 3.7 |
| Emergency ward | 12 | 5.6 |
| **Working experience** |  |  |
| <10 Years | 101 | 46.8 |
| 10 Years or more | 115 | 53.2 |
| **Current shift pattern** |  |  |
| Morning | 70 | 32.4 |
| Day | 104 | 48.1 |
| Evening | 17 | 7.9 |
| Night | 25 | 11.6 |

In of terms of working experience, slightly more than half of the respondents had 10 years or more of experience (115, 53.2%), while 101 (46.8%) had less than 10 years of experience. Regarding shift patterns, the largest share of participants were on day shifts (104, 48.1%), followed by those on morning shifts (70, 32.4%). Fewer respondents were on night shifts (25, 11.6%) and evening shifts (17, 7.9%) (Table 1).

### Level of Adherence to Hand Hygiene Practices

### Level of Adherence of Health Workers to HH Practices

**Figure 1:**
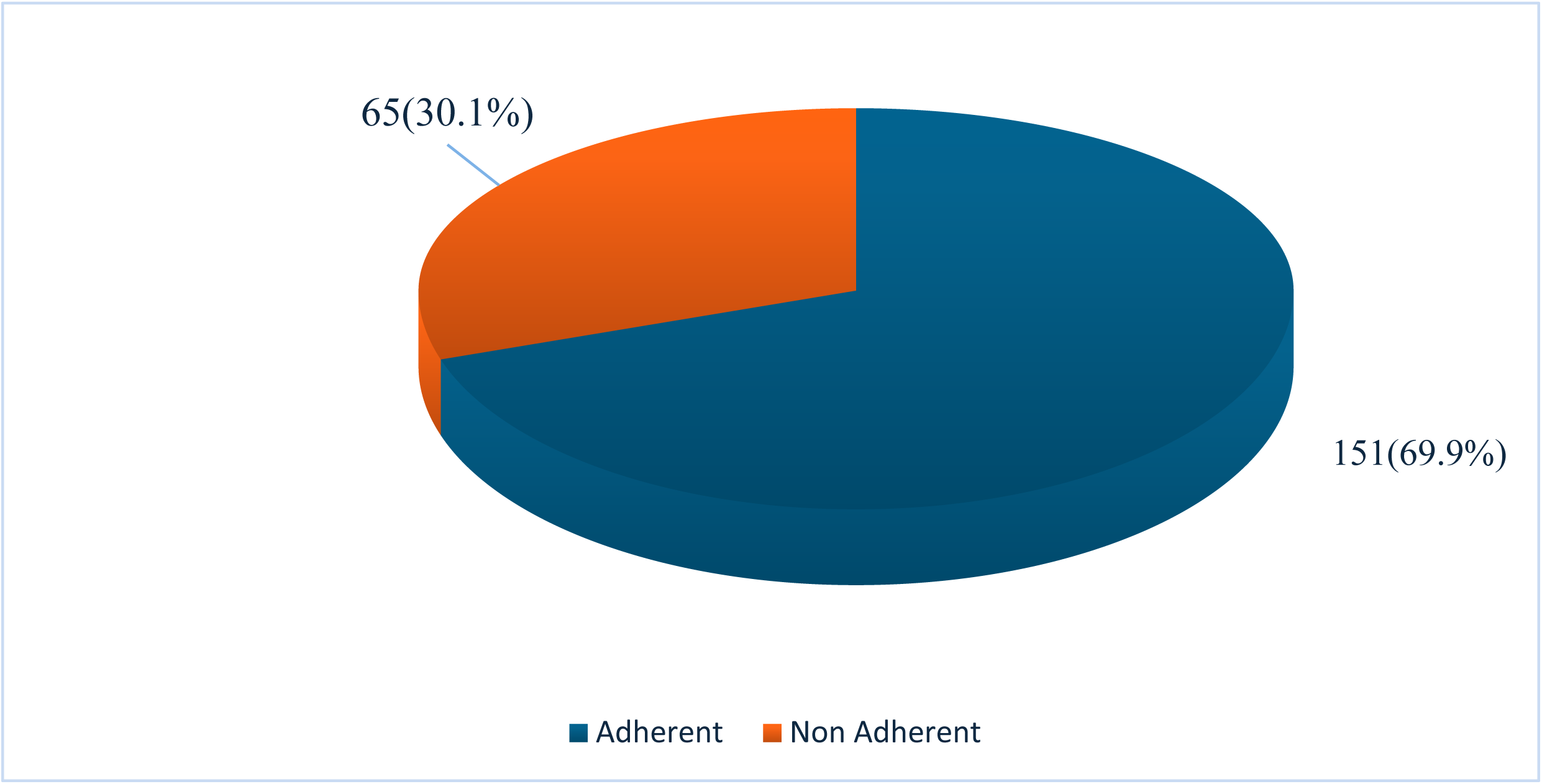
Adherence to Hand Hygiene Practices.

More than two-thirds of the participants, 151(69.9%) were adherent to hand hygiene practice (perform hand hygiene at ≥ 80% of required moments), while nearly one-third, 65(30.1%) were non-adherent (perform hand hygiene at < 80% of required moments) (Figure 1).

### Individual-Level Factors Associated with Adherence to Hand Hygiene Practices

The individual-level factors assessed in this study were: age, gender, cadre, working experience, knowledge-related factors, and attitude toward HH practices. Table 2 below shows the results according to the individual-level factors associated with adherence to HH practice Lubaga Hospital, Kampala City

**Table 2:** Individual-Level Factors Associated with Adherence to Hand Hygiene Practice.

| Factor | Adherence to Hand Hygiene Practices |  |  |  | cPR (95% CI); p-value | aPR (95% CI); p-value |
| --- | --- | --- | --- | --- | --- | --- |
|  | Adherent |  | Non Adherent |  |  |  |
|  | Freq. (N = 152) | Percent (%) | Freq. (N = 64) | Percent (%) |  |  |
| Age |  |  |  |  |  |  |
| <40 Years | 133 | 87.5 | 51 | 79.7 | 1.784(0.821-3.876); *p*=0.144 | 1.346(0.579-3.130); *p*=0.490 |
| 40 Years and above | 19 | 12.5 | 13 | 20.3 |  |  |
| Gender |  |  |  |  |  |  |
| Male | 19 | 12.5 | 15 | 23.4 | 0.467(0.220-0.990); *p*=0.047 | 0.490(0.226-0.969); *p*=0.048 |
| Female | 133 | 87.5 | 49 | 76.6 |  |  |
| **Cadre** |  |  |  |  |  |  |
| Nursing/Midwifery | 113 | 74.3 | 34 | 53.1 | **1.572(1.136-2.175); p=0.006** | **2.213(1.113-4.398); p=0.023** |
| Others | 39 | 25.7 | 30 | 46.9 |  |  |
| **Working experience** |  |  |  |  |  |  |
| <10 Years | 73 | 48 | 28 | 43.8 | 1.188(0.660-2.138); p=0.565 | 1.249(0.687-2.273); p=0.466 |
| 10 Years or more | 79 | 52 | 36 | 56.3 |  |  |
| **ABHR is recommended for most routine HH opportunities** |  |  |  |  |  |  |
| True | 98 | 64.5 | 47 | 73.4 | 0.656(0.344-1.253); p=0.202 | 0.681(0.351-1.323); p=0.257 |
| False/Not sure | 54 | 35.5 | 17 | 26.6 |  |  |
| **HH should be performed before aseptic procedures, even if gloves are worn** |  |  |  |  |  |  |
| True | 136 | 89.5 | 54 | 84.4 | 1.574(0.672-3.685); p=0.296 | 1.447(0.345-6.081); p=0.614 |
| False/Not sure | 16 | 10.5 | 10 | 15.6 |  |  |
| **Visibly soiled hands require soap and water, not ABHR** |  |  |  |  |  |  |
| True | 138 | 90.8 | 59 | 92.2 | 0.835(0.288-2.425); p=0.741 | 0.827(0.195-3.506); p=0.797 |
| False/Not sure | 14 | 9.2 | 5 | 7.8 |  |  |
| **Minimum rubbing time with ABHR is 20–30 seconds** |  |  |  |  |  |  |
| True | 129 | 84.9 | 52 | 81.3 | 1.294(0.600-2.792); p=0.511 | 1.599(0.660-3.871); p=0.298 |
| False/Not sure | 23 | 15.1 | 12 | 18.8 |  |  |
| **HH is required after touching patients' surroundings** |  |  |  |  |  |  |
| True | 145 | 95.4 | 62 | 96.9 | 0.668(0.135-3.308); p=0.621 | 0.533(0.064-4.426); p=0.560 |
| False/Not sure | 7 | 4.6 | 2 | 3.1 |  |  |
| **HH is recommended before donning gloves** |  |  |  |  |  |  |
| True | 127 | 83.6 | 52 | 81.3 | 1.172(0.548-2.507); p=0.682 | 1.791(0.741-4.327); p=0.195 |
| False/Not sure | 25 | 16.4 | 12 | 18.8 |  |  |
| **ABHR is more effective than soap for routine care** |  |  |  |  |  |  |
| True | 101 | 66.4 | 37 | 57.8 | 1.445(0.793-2.632); p=0.229 | 1.396(0.736-2.649); p=0.307 |
| False/Not sure | 51 | 33.6 | 27 | 42.2 |  |  |
| **HH reduces HAIs** |  |  |  |  |  |  |
| True | 137 | 90.1 | 60 | 93.8 | 0.609(0.194-1.911); p=0.395 | 0.418(0.112-1.553); p=0.193 |
| False/Not sure | 15 | 9.9 | 4 | 6.3 |  |  |
| **Wet hands reduce ABHR effectiveness** |  |  |  |  |  |  |
| True | 110 | 72.4 | 42 | 65.6 | 1.372(0.733-2.567); p=0.323 | 1.529(0.765-3.058); p=0.230 |
| False/Not sure | 42 | 27.6 | 22 | 34.4 |  |  |
| **Jewellery can reduce HH effectiveness** |  |  |  |  |  |  |
| True | 125 | 82.2 | 56 | 87.5 | 0.661(0.283-1.547); p=0.340 | 0.618(0.243-1.571); p=0.312 |
| False/Not sure | 27 | 17.8 | 8 | 12.5 |  |  |
| **Attitude toward HH Practices** |  |  |  |  |  |  |
| Good | 128 | 84.2 | 58 | 90.6 | **1.551(1.125-2.247); p=0.005** | **1.462(1.227-3.226); p=0.039** |
| Poor | 24 | 15.8 | 6 | 9.4 |  |  |

### Age

Age was not significantly associated with adherence in both crude and adjusted analyses (cPR = 1.784, 95% CI: 0.821–3.876, p = 0.144; aPR = 1.346, 95% CI: 0.579–3.130, p = 0.490). Accordingly, age was not associated with adherence to hand hygiene practices among participants (Table 2).

### Gender

Gender showed a statistically significant association with adherence (p≈0.038–0.048). Females demonstrated higher adherence (87.5%) compared to males (12.5% adherence vs 23.4% non-adherent among males). The crude prevalence ratio (cPR=0.467) and adjusted prevalence ratio (aPR=0.490) indicate that males were less likely to adhere to HH practices than females (Table 2).

### Cadre

Gender was significantly associated with adherence (cPR = 0.467, 95% CI: 0.220– 0.990, p = 0.047; aPR = 0.490, 95% CI: 0.226–0.969, p = 0.048). Accordingly, male participants were less likely to adhere to hand hygiene practices compared to female participants. (Table 2).

### Working Experience

There was no significant association between working experience and adherence (cPR = 1.188, 95% CI: 0.660–2.138, p = 0.565; aPR = 1.249, 95% CI: 0.687–2.273, p = 0.466), suggesting that level of experience was not associated with adherence to HH Practices (Table 2).

### Cadre

Cadre was significantly associated with adherence (cPR = 1.572, 95% CI: 1.136–2.175, p = 0.006; aPR = 2.213, 95% CI: 1.113–4.398, p = 0.023). As such, nursing and midwifery staff were significantly more likely to adhere to hand hygiene practices compared to other healthcare workers (Table 2).

### ABHR Recommendation for Routine Use

This variable was not significantly associated with adherence (cPR = 0.656, 95% CI: 0.344–1.253, p = 0.202; aPR = 0.681, 95% CI: 0.351–1.323, p = 0.257), indicating no relationship between this knowledge and adherence (Table 2).

### Hand Hygiene Before Aseptic Procedures

Knowledge of this was not significantly associated with adherence (cPR = 1.574, 95% CI: 0.672– 3.685, p = 0.296; aPR = 1.447, 95% CI: 0.345–6.081, p = 0.614) (Table 3).

### Use of Soap and Water for Soiled Hands

This variable was not significantly associated with adherence (cPR = 0.835, 95% CI: 0.288–2.425, p = 0.741; aPR = 0.827, 95% CI: 0.195–3.506, p = 0.797).

### Minimum ABHR Rubbing Time

Knowledge of this was not significantly associated with adherence (cPR = 1.294, 95% CI: 0.600– 2.792, p = 0.511; aPR = 1.599, 95% CI: 0.660–3.871, p = 0.298) (Table 2).

### Hand Hygiene After Patient Surroundings

This variable showed no significant association with adherence (cPR = 0.668, 95% CI: 0.135– 3.308, p = 0.621; aPR = 0.533, 95% CI: 0.064–4.426, p = 0.560) (Table 3).

### Hand Hygiene Before Donning Gloves

There was no significant association with adherence (cPR = 1.172, 95% CI: 0.548– 2.507, p = 0.682; aPR = 1.791, 95% CI: 0.741–4.327, p = 0.195) (Table 2).

### ABHR Effectiveness Compared to Soap

This variable was not significantly associated with adherence (cPR = 1.445, 95% CI: 0.793–2.632, p = 0.229; aPR = 1.396, 95% CI: 0.736–2.649, p = 0.307) (Table 2).

### Hand Hygiene and Reduction of HAIs

This was not significantly associated with adherence (cPR = 0.609, 95% CI: 0.194– 1.911, p = 0.395; aPR = 0.418, 95% CI: 0.112–1.553, p = 0.193) (Table 2).

### Effect of Wet Hands on ABHR Effectiveness

This factor was not significantly associated with adherence (cPR = 1.372, 95% CI: 0.733–2.567, p = 0.323; aPR = 1.529, 95% CI: 0.765–3.058, p = 0.230) (Table 2).

### Effect of Jewellery on Hand Hygiene

This variable was not significantly associated with adherence (cPR = 0.661, 95% CI: 0.283–1.547, p = 0.340; aPR = 0.618, 95% CI: 0.243–1.571, p = 0.312) (Table 2).

### Attitude Toward Hand Hygiene

Participants with a good attitude toward hand hygiene constituted 84.2% of adherent and 90.6% of non-adherent individuals. Attitude was significantly associated with adherence (cPR = 1.551, 95% CI: 1.125–2.247, p = 0.005; aPR = 1.462, 95% CI: 1.227–3.226, p = 0.039), an indication that respondents with a positive attitude were more likely to adhere to hand hygiene practices (Table 3).

### Availability of Relevant Resources and Adherence to Hand Hygiene Practice

The study assessed the association between availability of hand hygiene resources and adherence of health workers to hand hygiene practices at Lubaga Hospital, Kampala City.

**Table 3:** Availability of Relevant Resources and Adherence to HH Practice.

| Factor | Adherence to Hand Hygiene Practices |  |  |  | cPR (95% CI); p-value | aPR (95% CI); p-value |
| --- | --- | --- | --- | --- | --- | --- |
|  | Adherent |  | Non Adherent |  |  |  |
|  | Freq. (N = 152) | Percent (%) | Freq. (N = 64) | Percent (%) |  |  |
| ABHR sufficiently available at points of care |  |  |  |  |  |  |
| Yes | 80 | 52.6 | 34 | 53.1 | 1.072(0.748-1.538); *p*=0.704 | 1.027(0.120-4.308); *p*=0.371 |
| No | 72 | 47.4 | 30 | 46.9 |  |  |
| HH supplies are replenished promptly |  |  |  |  |  |  |
| Yes | 12 | 7.9 | 4 | 6.3 | 0.883(0.586-1.329);*p*=0.550 | 0.754(0.697-3.429); *p*=0.101 |
| No | 140 | 92.1 | 60 | 93.8 |  |  |

### Availability of Alcohol-Based Hand Rub (ABHR) at Points of Care

The crude analysis showed no significant association between ABHR availability and hand hygiene adherence (cPR = 1.072, 95% CI: 0.748–1.538, p = 0.704). Similarly, in the adjusted analysis, the association remained non-significant (aPR = 1.027, 95% CI: 0.120–4.308, p = 0.371). Accordingly, availability of ABHR at points of care was not significantly associated with adherence to hand hygiene practices among participants (Table 3).

### Timely Replenishment of Hand Hygiene Supplies

The crude prevalence ratio indicated no significant association (cPR = 0.883, 95% CI: 0.586– 1.329, p = 0.550). After adjustment, the association remained statistically non-significant (aPR = 0.754, 95% CI: 0.697–3.429, p = 0.101). Accordingly, prompt replenishment of hand hygiene supplies was not significantly associated with adherence to hand hygiene practices (Table 3).

### Organizational Factors Associated with Adherence to Hand Hygiene Practice

The study examined several organizational-level factors (namely, workload and patient load factors, training in HH and infection prevention, availability of visual reminders, leadership and organizational support, feedback and supervision, and teamwork and safety culture) influencing adherence to HH practices among healthcare workers at Lubaga Hospital.

### Average Number of Patients Per Shift

There was no statistically significant association between the number of patients per shift and adherence to hand hygiene practices in both crude and adjusted analyses (cPR = 0.926, 95% CI: 0.515–1.667, p = 0.926; aPR = 0.839, 95% CI: 0.460–1.533, p = 0.569), suggesting that workload in terms of patient volume was not significantly associated with adherence to hand hygiene practices (Table 4).

### Number of Direct Care Procedures Per Shift

This variables was not statistically significant with hand hygiene practices(cPR = 1.531, 95% CI: 0.851–2.754, p = 0.155; aPR = 1.724, 95% CI: 0.932–3.188, p = 0.083) (Table 4).

**Table 4:** Organizational Factors Associated with Adherence to Hand Hygiene Practice.

| Factor | Adherence to Hand Hygiene Practices |  |  |  | cPR (95% CI); p-value | aPR (95% CI); p-value |
| --- | --- | --- | --- | --- | --- | --- |
|  | Adherent |  | Non Adherent |  |  |  |
|  | Freq. (N = 152) | Percent (%) | Freq. (N = 64) | Percent (%) |  |  |
| **Average number of patients per shift in the past week** |  |  |  |  |  |  |
| Less than 100 | 66 | 43.4 | 29 | 45.3 | 0.926(0.515-1.667); p=0.926 | 0.839(0.460-1.533); p=0.569 |
| 100 and above | 86 | 56.6 | 35 | 54.7 |  |  |
| **Number of direct care procedures per shift** |  |  |  |  |  |  |
| <10 | 85 | 55.9 | 29 | 45.3 | 1.531(0.851-2.754); p=0.155 | 1.724(0.932-3.188); p=0.083 |
| 10 or more | 67 | 44.1 | 35 | 54.7 |  |  |
| **Self-rated workload** |  |  |  |  |  |  |
| Very low/Low | 79 | 52 | 38 | 59.4 | 0.740(0.410-1.338); p=0.320 | 0.650(0.351-1.203); p=0.170 |
| Moderate/High | 73 | 48 | 26 | 40.6 |  |  |
| **Received formal HH/IPC training in the past 12 months** |  |  |  |  |  |  |
| Yes | 89 | 58.6 | 38 | 59.4 | 1.967(1.534-2.758); p<0.001 | 1.771(1.673-2.989); p=0.001 |
| No | 63 | 41.4 | 26 | 40.6 |  |  |
| **When formal training in HH/IPC was received** |  |  |  |  |  |  |
| <12 months back | 62 | 40.8 | 28 | 43.8 | 0.886(0.491-1.598); p=0.687 | 0.768(0.414-1.425); p=0.403 |
| 12 or more months ago/Not Applicable | 90 | 59.2 | 36 | 56.3 |  |  |
| **Posters/reminders are visible and helpful** |  |  |  |  |  |  |
| Yes | 13 | 8.6 | 5 | 7.8 | 1.945(1.667-5.339); p=0.028 | 1.747(1.538-5.949); p=0.042 |
| No | 139 | 91.4 | 59 | 92.2 |  |  |
| **Unit leadership supports HH** |  |  |  |  |  |  |
| Yes | 129 | 84.9 | 53 | 82.8 | 0.932(0.671-1.296); p=0.677 | 0.768(0.414-1.425); p=0.403 |
| No | 23 | 15.1 | 11 | 17.2 |  |  |
| **Receive feedback on HH performance** |  |  |  |  |  |  |
| Yes | 132 | 86.8 | 50 | 78.1 | 0.704(0.512-0.968); p=0.031 | 0.339(0.156-0.738); p=0.006 |
| No | 20 | 13.2 | 14 | 21.9 |  |  |
| **HH supervision frequency** |  |  |  |  |  |  |
| Daily | 111 | 73 | 42 | 65.6 | 1.418(0.757-2.657); p=0.276 | 1.871(0.575-6.090); p=0.298 |
| Not Daily | 41 | 27 | 22 | 34.4 |  |  |
| **Staff freely speak up about infection risks** |  |  |  |  |  |  |
| Yes | 143 | 94.1 | 63 | 98.4 | 1.834(0.906-3.712); p=0.092 | 1.856(0.815-4.230); p=0.141 |
| No | 9 | 5.9 | 1 | 1.6 |  |  |
| **Management acts on safety concerns** |  |  |  |  |  |  |
| Yes | 143 | 94.1 | 55 | 85.9 | 1.639(0.908-2.961); p=0.101 | 1.874(0.203-3.767); p=0.857 |
| No | 9 | 5.9 | 9 | 14.1 |  |  |
| **Teamwork supports safe care** |  |  |  |  |  |  |
| Yes | 129 | 84.9 | 56 | 87.5 | 1.444(1.007-2.068); p=0.045 | 3.006(1.424-6.348); p=0.004 |
| No | 23 | 15.1 | 8 | 12.5 |  |  |

### Self-Rated Workload

There was no significant association between self-rated workload and adherence to hand hygiene practices (cPR = 0.740, 95% CI: 0.410–1.338, p = 0.320; aPR = 0.650, 95% CI: 0.351–1.203, p = 0.170). Accordingly, perceived workload did not significantly affect hand hygiene adherence (Table 4).

### Received formal HH/IPC training in the past 12 months

This factor was strongly and significantly associated with adherence to hand hygiene practices in both crude and adjusted analyses (cPR = 1.967, 95% CI: 1.534–2.758, p < 0.001; aPR = 1.771, 95% CI: 1.673–2.989, p = 0.001). Those who received formal HH/IPC training were significantly more likely to adhere to hand hygiene practices (Table 4).

### Timing of formal HH/IPC training

This variable was not significantly associated with adherence to hand hygiene practices (cPR = 0.886, 95% CI: 0.491–1.598, p = 0.687; aPR = 0.768, 95% CI: 0.414–1.425, p = 0.403), an indication that timing of training is not significantly associated with adherence to hand hygiene practices (Table 4).

### Posters/Reminders Visibility and Usefulness

This factor was significantly associated with adherence (cPR = 1.945, 95% CI: 1.667– 5.339, p = 0.028; aPR = 1.747, 95% CI: 1.538–5.949, p = 0.042). Availability of visible reminders was associated with improved adherence, although the small numbers suggest this should be interpreted cautiously (Table 4).

### Unit Leadership Support for Hand Hygiene

Most participants reported that unit leadership supported hand hygiene (84.9% adherent and 82.8% non-adherent). However, no significant association was observed (cPR = 0.932, 95% CI: 0.671– 1.296, p = 0.677; aPR = 0.768, 95% CI: 0.414–1.425, p = 0.403). Accordingly, leadership support was not significantly associated with adherence to HH Practice in this study (Table 4).

### Feedback on Hand Hygiene Performance

This variable showed a significant association in both crude and adjusted analyses (cPR = 0.704, 95% CI: 0.512–0.968, p = 0.031; aPR = 0.339, 95% CI: 0.156–0.738, p = 0.006). Accordingly, receiving feedback significantly improved adherence to hand hygiene practices (Table 4).

### Hand Hygiene Supervision Frequency

This variable was not significantly associated with adherence (cPR = 1.418, 95% CI: 0.757–2.657, p = 0.276; aPR = 1.871, 95% CI: 0.575–6.090, p = 0.298). Accordingly, frequency of supervision did not significantly affect adherence to hand hygiene practices (Table 4).

### Staff Speaking Up About Infection Risks

There was no statistically significant association with adherence (cPR = 1.834, 95% CI: 0.906– 3.712, p = 0.092; aPR = 1.856, 95% CI: 0.815–4.230, p = 0.141). Accordingly, speaking up culture was not significantly associated with adherence to hand hygiene practices (Table 4).

### Management Action on Safety Concerns

This variable was not significantly associated with adherence (cPR = 1.639, 95% CI: 0.908–2.961, p = 0.101; aPR = 1.874, 95% CI: 0.203–3.767, p = 0.857). Accordingly, management action on safety concerns was not significantly associated with adherence to hand hygiene practices (Table 4).

### Teamwork Supporting Safe Care

This variable was significantly associated with adherence in both crude and adjusted analyses (cPR = 1.444, 95% CI: 1.007–2.068, p = 0.045; aPR = 3.006, 95% CI: 1.424–6.348, p = 0.004). Accordingly, strong teamwork was a key predictor of adherence, with participants being about three times more likely to adhere when teamwork supported safe care (Table 4).

## Discussion

### Level of Adherence to Hand Hygiene Practices

This study found that 69.9% of healthcare workers were adherent to hand hygiene (HH) practices. This relatively high adherence level may be attributed to increased awareness of infection prevention and control (IPC), especially following global and national emphasis during outbreaks such as COVID-19. This finding is higher than reports from several Ugandan and East African studies, where adherence levels ranged between 30% and 60% [10]. However, it is consistent with studies conducted in better-resourced or training-focused settings, where adherence levels above 65% have been reported [11]. Conversely, some high-income settings report adherence exceeding 80%, indicating that while Lubaga Hospital performs relatively well, there is still a gap compared to optimal standards.

Accordingly, while progress has been made, nearly one-third of healthcare workers remain non-adherent, posing a continued risk for healthcare-associated infections (HAIs). These findings imply that sustained interventions are needed to further improve adherence to HH practices expected of healthcare professionals. Without this, the health facility might experience HIAs, along with associated negative consequences, such as increased healthcare costs.

### Individual-Level Factors Associated with Adherence to Hand Hygiene Practice Gender

Female healthcare workers were significantly more likely to adhere to HH practices than males. This may be explained by differences in risk perception, hygiene behavior, and compliance tendencies, as females are often reported to demonstrate more health-conscious behaviors. This finding agrees with studies from Kenya and Ethiopia, which found higher HH compliance among female workers [12, 13]. However, some studies in Europe have found no gender differences, suggesting that cultural and contextual factors may be linked to this relationship [14]. This finding implies that gender-responsive strategies may be needed, particularly targeting male healthcare workers to improve adherence.

### Cadre

Nurses and midwives were more likely to adhere compared to other cadres. This is likely due to their frequent patient contact, structured IPC training, and strict nursing protocols. This aligns with findings from Uganda and Tanzania, where nurses exhibited higher compliance due to continuous bedside care responsibilities [15]. However, some studies in high-income countries show minimal cadre differences due to standardized protocols across all professions [16]. This finding implies that non-nursing cadres may require targeted training and accountability systems to improve adherence.

### Attitude toward HH Practices

A positive attitude was strongly associated with adherence. This suggests that beliefs, motivation, and perception of the importance of HH are critical determinants of behavior. This is consistent with studies globally showing that attitudinal factors outweigh knowledge in predicting compliance [17]. In contrast, some studies in resource-limited settings have found knowledge to play a stronger role, indicating contextual differences [18]. This finding implies that behavioral change interventions should prioritize attitude transformation rather than knowledge alone.

### Availability of Relevant Resources and Adherence to Hand Hygiene Practices

The study found no significant association between the availability of ABHR or supply replenishment and adherence. This may be due to uniform distribution of resources across the facility, limiting variation, or due to behavioral factors outweighing structural ones. This finding contrasts with WHO evidence that improved access significantly increases compliance [19–21]. However, it aligns with studies from Uganda showing that availability alone is insufficient without behavioral reinforcement [22–23].

### Organizational Factors Associated with Adherence to Hand Hygiene Practice Training in HH/IPC

Healthcare workers who received training were significantly more likely to adhere. This may be because training reinforces knowledge, builds skills, and increases awareness of infection risks. This agrees with findings from Rwanda [24] and Uganda [4, 10], where training doubled compliance rates. However, the timing of training was not significant, suggesting that continuous reinforcement may be more important than frequency [25,26]. This, therefore, calls for the need for ongoing training programs in order to improve healthcare workers’ adherence to HH practices.

### Visual Reminders

Posters/reminders significantly improved adherence. These act as constant cues that trigger appropriate behavior at the point of care. This aligns with WHO guidelines emphasizing reminders as a core strategy [19. Similar findings have been reported in Ethiopia and Kenya [12].

### Feedback on HH Performance

Receiving feedback significantly improved adherence. This may be due to increased accountability and awareness of performance gaps. This is consistent with global evidence showing that audit and feedback interventions significantly improve compliance [21].

### Teamwork and Safety Culture

Strong teamwork significantly improved adherence. This suggests that peer influence, shared responsibility, and supportive environments promote compliance. This agrees with studies showing that positive safety culture is a key predictor of HH compliance [16, 27–29].

## Conclusion

The study established that adherence to hand hygiene practices among healthcare workers at Lubaga Hospital was moderately high at 69.9%; however, this level remains suboptimal when compared to recommended infection prevention and control standards. The findings demonstrate that adherence is significantly shaped by both individual and organizational dynamics. Specifically, gender, professional cadre, and attitude toward hand hygiene were key individual determinants, while organizational factors such as training, availability of visual reminders, performance feedback, and teamwork significantly enhanced compliance.

In contrast, structural and resource-related factors, including the availability of hand hygiene supplies and workload indicators, were not significantly associated with adherence, suggesting that the mere presence of resources does not guarantee appropriate practice. Overall, this study underscores that behavioral, cultural, and system-level linkages, particularly those that reinforce motivation, accountability, and teamwork, are more critical in driving hand hygiene compliance than infrastructural provisions alone. Therefore, effective interventions must prioritize integrated strategies that combine behavioral change approaches with supportive institutional frameworks to achieve optimal and sustained adherence to hand hygiene practices.

## Data Availability

Data will be availed on request The minimal data set is available

## Acknowledgements

Gratitude goes out to my Supervisor and the Biostatistician. Special appreciation is extended to the management and staff of Lubaga Hospital for their cooperation and support during data collection. Finally, heartfelt appreciation to family, friends and colleagues whose encouragement and support made this research possible.

## Ethical approval

Ethical considerations involved approval of the research proposal by the administration of the faculty of health sciences of Uganda Martyrs University, followed by ethical approval by the Research Ethics Committee (REC) of Mildmay Uganda, and administrative clearance from Lubaga Hospital, Kampala. The study respondents gave informed consent before being enrolled to participate in the study. They were informed that participation in the study is at free will and that they are at liberty to withdraw their participation at any stage. Anonymity was maintained using codes as opposed to respondents’ specific identifiers. Permission for access to electronic data was obtained from the Ministry of Health Assistant Commissioner in charge of Health Information Management and the director of Lubaga Hospital, Kampala.

## Consent for publication

Study participants gave informed consent for publishing the study outcomes.

## Availability of data and materials

All the data analyzed during this study are included in this manuscript. The data sets generated and analyzed during the study are also available from the corresponding author on request.

## Author’s Contributions

NF conceptualized and designed the study, coordinated data collection, performed statistical analysis, interpreted the findings, and drafted the manuscript. KO contributed to the study design, provided methodological guidance, supervised the research process, and critically reviewed the manuscript for intellectual content.

## Supporting information

### S1: REC APPROVAL

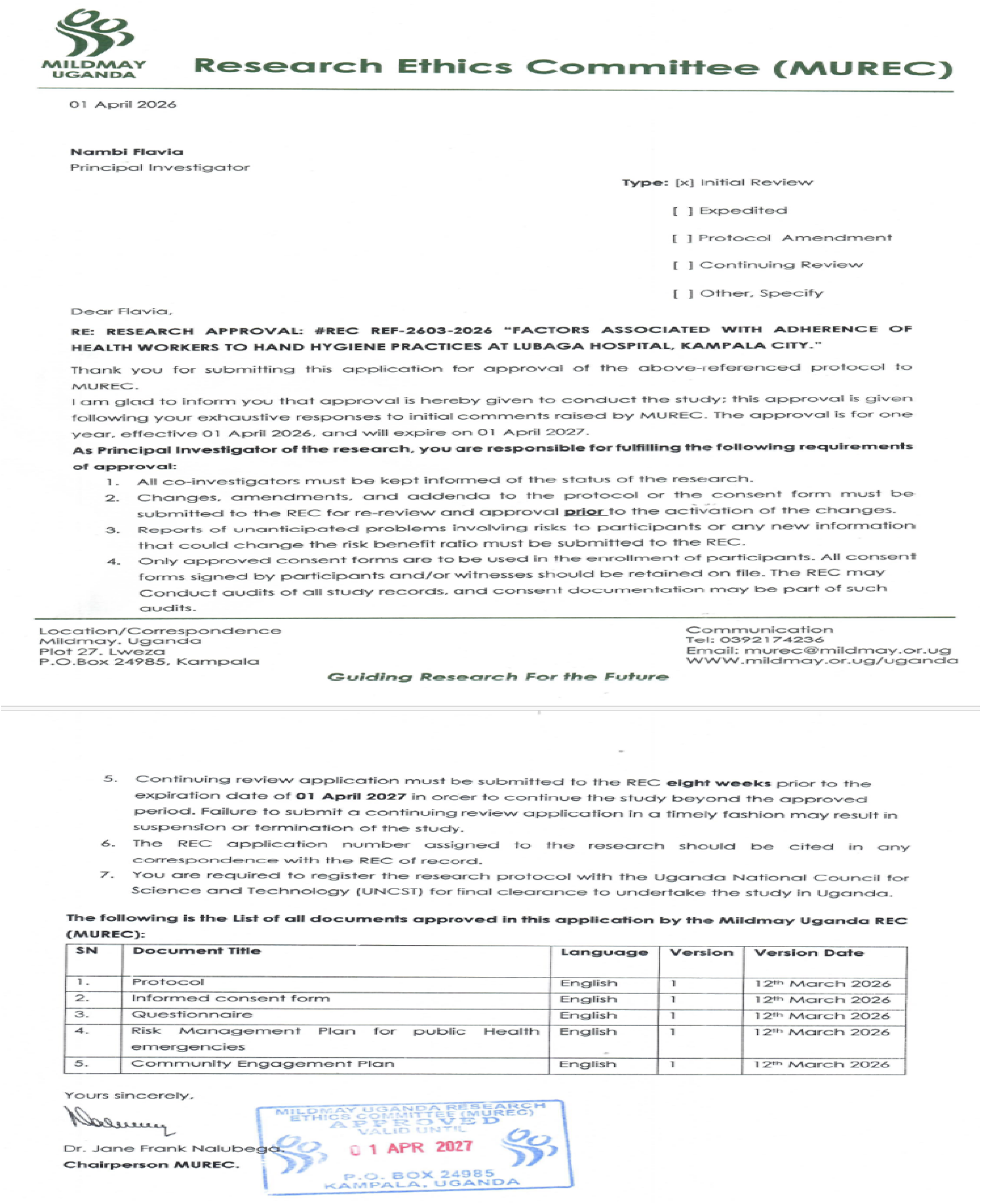

### S2. Institutional Approval For the Study

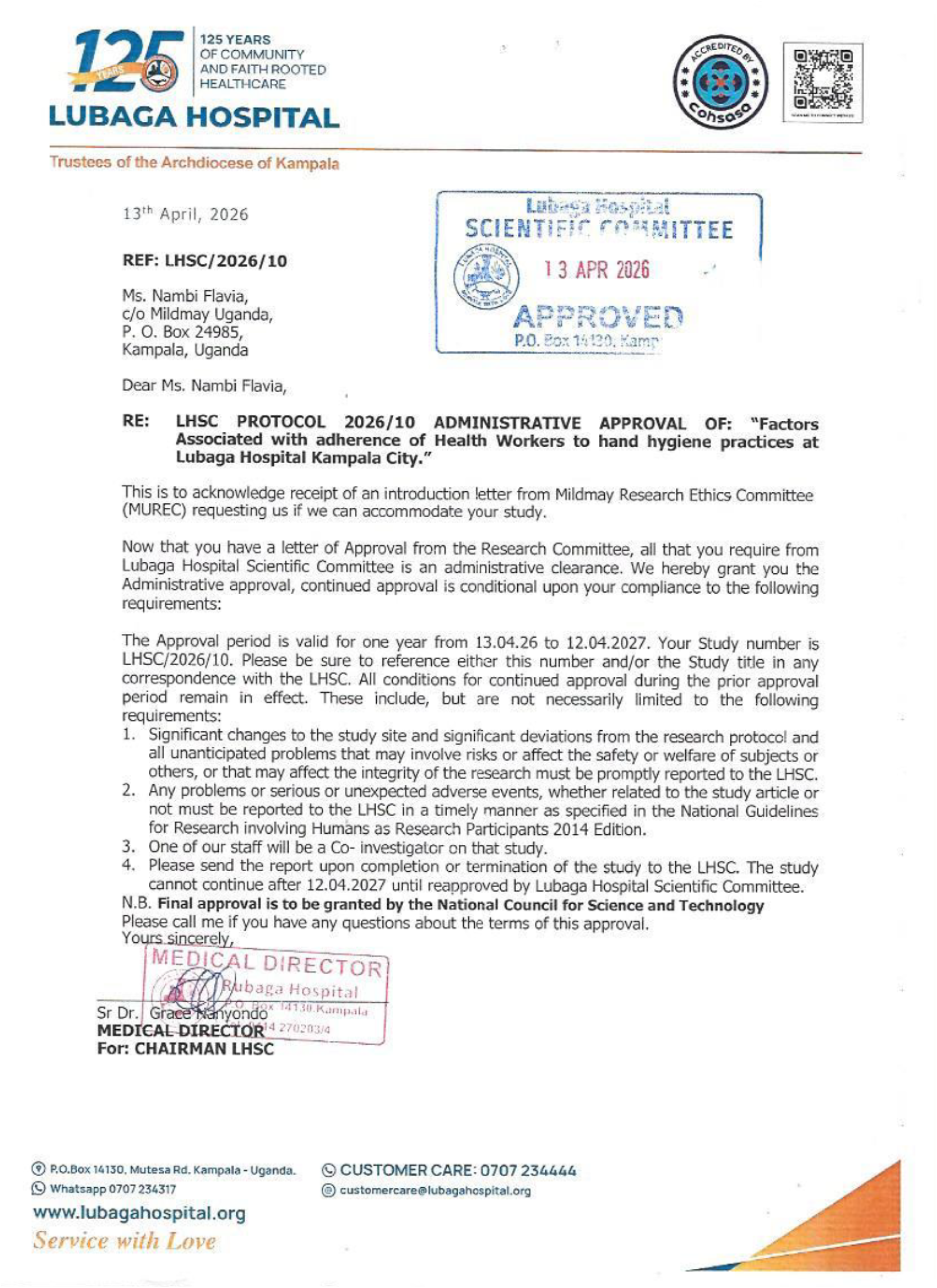

